# Transcranial photobiomodulation rebalances cortical excitation/inhibition in young adults with attention deficit/hyperactivity disorder

**DOI:** 10.64898/2026.09.01.26361884

**Authors:** Xuye Yuan, Yiyang Wang, Chen Dang, Hongyu Liu, Lili Yang, Dongwei Li, Li Sun, Yan Song

## Abstract

**Background:** Attention deficit/hyperactivity disorder (ADHD) is a neurodevelopmental condition lacking mechanistically grounded interventions. Here, we tested whether transcranial photobiomodulation (tPBM) can restore neural homeostasis in ADHD patients.

**Methods:** In a randomized, double-blind, sham-controlled crossover design, 28 young adults with ADHD completed a two-week intervention, receiving active (150 mW) and sham (0 mW) stimulation over the right prefrontal cortex for 16 minutes with concurrent electroencephalography (EEG) recording, alongside 29 healthy controls providing a normative reference.

**Results:** Behaviorally, tPBM improved working memory K scores in the ADHD group, with performance closer to typical levels. Across sensor and source levels, tPBM progressively increased relative alpha power, steepened the aperiodic exponent, and enhanced neural complexity, as indexed by multiscale entropy, with widespread effects spanning frontoparietal and attention systems, extending to sensory and default-mode regions, collectively indicating a shift toward normative neural dynamics. Notably, these changes—consistent with rebalanced excitation/inhibition dynamics—predict behavioral improvements in working memory.

**Conclusions:** Together, our findings identify tPBM as a candidate approach for restoring excitation/inhibition balance and normalizing large-scale neural dynamics in ADHD patients, providing a mechanistic foundation for its therapeutic potential.

## 1. Introduction

Attention-deficit/hyperactivity disorder (ADHD) is a prevalent neurodevelopmental disorder affecting millions of individuals worldwide [1]. It is characterized by developmentally inappropriate levels of inattention, disorganization, and hyperactivity–impulsivity, often accompanied by deficits in working memory that contribute to its core symptoms [2,3]. Once regarded solely as a childhood condition, ADHD is now recognized in approximately 3–5% of adults, reflecting its enduring impact later in life [4]. Confronted with the developmental and psychosocial challenges of early adulthood, young adults with ADHD often encounter pronounced difficulties in academic, occupational, and social functioning [5,6]. Although the diagnosis and intervention of adult ADHD have attracted increasing attention, the quest for more effective, evidence-guided interventions remains a critical challenge.

Previous studies have indicated that an optimal excitatory/inhibitory (E/I) balance is critical for the functionality of the brain, and in ADHD, this balance is often shifted toward excitation (E > I) [7]. Animal studies have shown that mice with gene deletions of actin-regulating proteins exhibit excessive striatal glutamate signaling, leading to heightened excitation and substantial ADHD-like hyperactivity, impulsivity, and working memory deficits [8,9]. In humans, electroencephalography (EEG) recordings reveal altered spectral dynamics [10,11], reflecting weakened inhibitory control, alongside biochemical reports of increased excitatory metabolites in the anterior cingulate cortex and broader disruptions in gamma-aminobutyric acid (GABA)-mediated inhibition [12,13]. These findings suggest a functional role for cortical E/I dysregulation in ADHD pathology and motivate intervention approaches capable of restoring neural balance [14].

Oscillatory rhythms reflect the balance between excitatory and inhibitory activity, with increased alpha power indicating reduced excitability and greater cortical synchronization [15,16]. In parallel, aperiodic activity reflects the synaptic-level E/I balance, with flatter spectral exponents reflecting increased excitability and reduced inhibition [17]. From an information-coding perspective, both components are related to neural synchrony and informational capacity [18]. Beyond spectral features, multiscale entropy (MSE) quantifies the temporal complexity of neural activity across multiple timescales [19], which is thought to peak under optimal E/I balance [20,21]. In ADHD, E/I dysregulation emerges across oscillatory, aperiodic, and network-level dynamics [11,22–24]. These complementary features provide a multidimensional view of cortical function, offering insight into the E/I-modulated fine-grained structure of neural dynamics.

Transcranial photobiomodulation (tPBM) is a safe, noninvasive neuromodulation technique that delivers near-infrared light to the cortex and enhances neuronal activity through mitochondrial metabolism via cytochrome *c* oxidase [25–27]. Beyond bioenergetic effects, evidence suggests that tPBM may modulate cortical excitability through E/I-related mechanisms. Preclinical studies have shown that tPBM can reduce excessive excitation by enhancing ATP-dependent sodium-potassium pump activity and stabilizing membrane potential, limiting pathological depolarization [28,29]. Evidence from healthy adults indicates that tPBM modulates intrinsic brain activity related to inhibition, including increases in EEG alpha power across large-scale networks such as the default mode, executive control, frontoparietal, and visual networks, and transiently reduces cortical excitability, as reflected by decreased motor-evoked potential [30–33]. These mechanistic effects further support the promise of tPBM in individuals with ADHD.

Early work in ADHD animal models revealed that tPBM can decrease hyperactivity and preserve neural integrity [34]. A small case series echoes this potential, describing rapid ADHD symptom relief following a single session of stimulation using combined 905 nm superpulsed and 660 nm lasers without medication [35]. Moreover, a study in healthy young adults demonstrated that 1064-nm tPBM applied to the right prefrontal cortex (rPFC) enhances visual working memory capacity, accompanied by increased contralateral delay activity (CDA), paving the way for targeting core cognitive deficits in ADHD [36]. Recently, a behavioral study in adults with ADHD revealed that a multisession tPBM protocol improved working memory and sustained attention, with effects lasting up to four weeks [37]. Despite these findings, the neurophysiological mechanisms underlying the effects of tPBM on ADHD remain insufficiently understood, highlighting the need for a systematic investigation of how tPBM modulates neural dynamics to support cognition.

Emerging evidence increasingly conceptualizes ADHD as a disorder of large-scale brain network organization, with atypical cortical E/I dynamics potentially contributing to widespread abnormalities and functional disorders [38,39]. To this end, we employed a randomized, double-blind, sham-controlled design to test whether tPBM modulates neural markers of cortical E/I imbalance in young adults with ADHD, including alpha oscillations, aperiodic slope, and MSE, while further delineating its effects across large-scale cortical systems and linking these changes to their core cognitive deficits.

## 2. Methods and materials

### 2.1. Participants

Participants with ADHD were recruited through the Peking University Sixth Hospital/Institute of Mental Health. Healthy controls (HCs) were recruited through posters and social media. The study protocol received approval from the hospital’s Ethics Committee (approval code: 2021-46) and was registered with the Chinese Clinical Trial Registry (ChiCTR2500108136). Written informed consent was obtained from all participants, and all procedures adhered to the Declaration of Helsinki.

All participants underwent a psychiatric evaluation by an experienced psychiatrist. Individuals in the ADHD group met the diagnostic criteria for adult ADHD according to the *Diagnostic and Statistical Manual of Mental Disorders, Fifth Edition* (DSM-V) [40], whereas HCs did not. All participants completed the Adult ADHD Self-Report Scale (ASRS) to assess ADHD symptoms [41]. To reduce potential confounds, eligibility was restricted to individuals who met the following criteria: (1) no prior head injury involving loss of consciousness; (2) absence of current psychiatric disorders, as assessed using the *Mini International Neuropsychiatric Interview* (MINI) [42], including mood, anxiety, and psychotic disorders; (3) no evidence of intellectual disability based on clinical or educational records; and (4) normal or corrected-to-normal vision, with no color vision deficiency.

In total, 43 individuals with ADHD were assessed for eligibility. Thirty-three participants were enrolled, of whom 2 withdrew, leaving 31 completers. In parallel, 33 HCs were recruited. Overall, the final analyses included 28 participants with ADHD and 29 HCs (Fig. 1A). Symptom assessments confirmed that ADHD participants exhibited significantly elevated inattention and hyperactivity/impulsivity scores relative to those of HCs (Table 1).

**Fig. 1.**
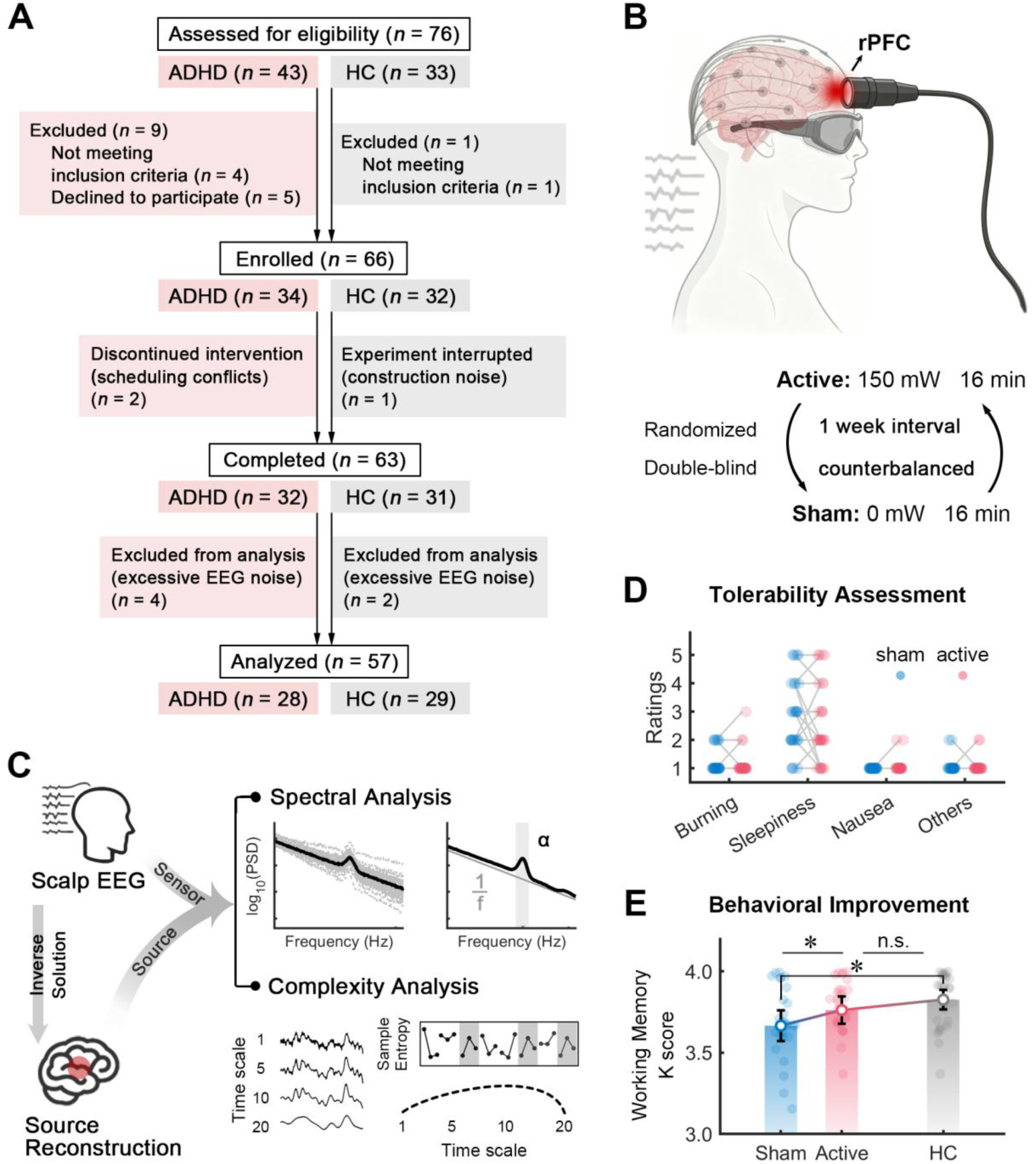
Study design, intervention protocol, and behavioral assessment. **(A)** CONSORT flow diagram illustrating participant recruitment, exclusion, and final inclusion in the analysis. **(B)** Stimulation procedure showing a 16-minute tPBM targeting the rPFC in both active and sham conditions under a counterbalanced design. **(C)** Simultaneous EEG recording during the intervention, with data analyzed at both the sensor and source levels using spectral and complexity measures to characterize tPBM-related neural dynamics. **(D)** Self-reported tolerability of tPBM in the ADHD group. Ratings of burning, sleepiness, nausea, and other unpleasant feelings did not differ between active and sham conditions. **(E)** Compared with that in HCs, behavioral performance in working memory, quantified using the K score, was lower in the ADHD group under the sham condition and improved following active tPBM, eliminating group differences. n.s., not significant difference, \**p*_FDR_ < 0.05.

**Table 1.** Demographic and clinical characteristics of the participants.

| Characteristics | ADHD (*n* = 28) | HC (*n* =29) | $\chi^2 / t$ | *p* |
| --- | --- | --- | --- | --- |
| Sex (male: female) | 14: 14 | 15: 14 | 0.017 | 0.896 |
| Age (years) | 24.752 ± 4.805 | 23.470 ± 2.682 | 1.249 | 0.217 |
| Symptom |  |  |  |  |
| ASRS<sub>inattention</sub> | 25.036 ± 3.844 | 8.931 ± 4.183 | 15.120 | < 0.001 |
| ASRS<sub>hyperactivity/impulsivity</sub> | 20.500 ± 5.081 | 8.103 ± 4.665 | 9.610 | < 0.001 |

The values are the means ± standard deviations. ADHD, attention deficit/hyperactivity disorder; ASRS, adult ADHD self-report scale; HC, healthy control.

Additionally, 23 participants with ADHD had a history of medication use. To limit potential influences, their stimulant medication was temporarily paused for at least one week before testing, following clinical recommendations and with the oversight of their treating clinicians. All data were collected prior to any medication intake, and participants returned to their usual treatment regimen after completing the study.

### 2.2. tPBM Procedure

A diode-pumped solid-state laser (1064 ± 1 nm; Jieliang Medical Device Inc., Jiangxi, China) delivered stimulation through a uniform 13.57 cm^2^ beam (4 cm diameter) at a continuous output of 2036 mW, corresponding to an irradiance of 150 mW/cm². The selected parameters complied with ANSI laser safety standards and have shown no adverse effects previously [36,43]. Stimulation was administered over the rPFC (FP2) using a handheld applicator (Fig. 1B). Hair was parted to reduce attenuation, and participants sat comfortably in a dimly lit room, remained awake with eyes open, and maintained stable device contact. Both participants and experimenters wore protective goggles.

Each condition lasted 960 s and consisted of either active stimulation (150 mW/cm²) or sham stimulation (0 mW/cm²). Participants received both conditions in a counterbalanced order separated by at least one week to minimize carryover effects (Fig. 1B). HCs completed a no-stimulation condition to provide a matched baseline. During the sham condition, the device remained powered with the laser output set to 0 mW so that operational sound and handling were preserved, reducing expectancy bias. Immediately afterward, participants completed a sensation questionnaire assessing potential tPBM-related experiences (sensations of burning, sleepiness, nausea, and other discomforts) and performed a validated working memory task [36]. Responses were rated on a 5-point Likert scale ranging from 1 (no sensation) to 5 (maximum).

### 2.3. EEG Recording and Preprocessing

Resting-state EEGs were acquired throughout the stimulation period using a 64-channel SynAmps system (NeuroScan Inc.) arranged according to the international 10–20 system (Fig. 1C). The signals were referenced online to the left mastoid, sampled at 500 Hz, and band-limited between 0.01 and 200 Hz. Electrode impedance was kept below 5 kΩ. Vertical EOG was recorded from electrodes placed above and below the left eye, and horizontal EOG was recorded from electrodes at the outer canthi.

Data preprocessing was performed in MATLAB (MathWorks, Natick, MA) using EEGLAB [44]. The signals were downsampled to 250 Hz and bandpass filtered between 0.1 and 40 Hz, and bad channels were interpolated before they were rereferenced to the average of all electrodes. The 960-s stimulation interval was divided into four 240-s segments, and each segment was further segmented into 2-s epochs with a 1-s overlap. Epochs containing obvious artifacts were first removed manually. Independent component analysis (ICA) was then applied to identify and remove components associated with ocular, muscular, and cardiac activity. Finally, epochs were automatically rejected when the voltage fluctuations exceeded ±80 μV at any electrode. Retention rates were 90.01% for ADHD-active conditions, 90.93% for ADHD-sham conditions, and 91.66% for HCs and did not differ across stages, stimulation conditions, or groups (Supplementary Fig. 1).

### 2.4. Behavioral Analysis

Working memory was selected as a primary outcome measure in ADHD and was tested immediately after the intervention. We examined participants’ responses in the visual working memory task using K scores following previous study [36]. K scores were computed for each memory load and subsequently averaged across loads to obtain an overall measure of working memory performance for each participant.

### 2.5. Spectral Analysis

The power spectral density (PSD) for each channel was estimated using Welch’s method with a 2-s window and 50% overlap. The resulting PSDs were then parameterized using the FOOOF algorithm to decompose the spectrum into aperiodic and periodic components [17]. The algorithm was applied to spectral data in the 3–40 Hz range with the following settings: peak width limit = [1,6], maximum number of peaks = 6, minimum peak height = 0.05, peak threshold = 1.5, and aperiodic mode = ‘fixed’. From the aperiodic component, the exponent of the 1/*f*-like slope was extracted for further analysis. For the periodic component, the oscillatory power was computed after the aperiodic contribution was removed. To account for interindividual differences, relative alpha power (8–12 Hz) was obtained by normalizing power in the alpha band to the total power of the periodic component [45].

### 2.6. Complexity Analysis

Signal complexity and irregularity in the EEG were quantified with MSE, which estimates sample entropy (SampEn) across 20 time scales through a coarse-graining procedure, with larger scales corresponding to progressively lower sampling rates [19]. SampEn is defined as the negative natural logarithm of the probability that sequences of length *m* remain similar at the next time point as follows:

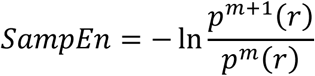

where the sequence length *m* is set to 2 and the similarity criterion *r* (the tolerance within which two points are considered similar) is set to 0.5. Owing to the scale-dependent changes in signal variation, *r* was recalculated at each time scale [46].

### 2.7. Source Reconstruction

Source reconstruction was performed using the Brainstorm Toolbox [47]. A realistic three-layer (scalp, skull, and brain) boundary element model (BEM) of the head was constructed on the basis of the ICBM152 template anatomy, with electrodes coregistered to the scalp surface. The lead-field matrix was computed using the OpenMEEG plugin, and the weighted minimum norm estimate (MNE) was applied to project sensor-space EEG onto 5,000 cortical source vertices. Source time series were parcellated according to the Schaefer atlas [48], with 100 regions of interest (ROIs) assigned to the Yeo 7 networks [49], and the dominant pattern across all vertices within each ROI was extracted via principal component analysis (PCA). Spectral and complexity analyses were then applied to these ROI-level signals, using the same procedures as for the sensor-level data.

### 2.8. Normative Template-Based Analysis

A normative reference was constructed from HC participants by averaging feature values across individuals to obtain a baseline template. The Mahalanobis distance between each participant and the HC template was then calculated using a covariance matrix estimated from the HC data with a Ledoit–Wolf shrinkage estimator [50,51]. Distance values were computed separately for the active and sham conditions to assess deviations from the HC reference, with lower values indicating greater similarity to the normative profile. A reduction in distance in the active condition relative to the sham condition indicated that tPBM shifted individual brain activity toward a more normative (HC-like) pattern.

### 2.9. Statistical Analysis

Parametric statistical analyses were performed in JASP [52]. Within the ADHD group, differences between the active and sham conditions were assessed using paired-sample *t* tests for tolerability and K scores. Independent-sample *t* tests compared K scores between the ADHD group and the HC group under each condition with false discovery rate (FDR) correction.

For EEG measures (alpha power, aperiodic exponent, and multiscale entropy), cluster-based permutation tests (5,000 Monte Carlo iterations; maxsum criterion) were implemented in the FieldTrip toolbox to compare active and sham conditions within the ADHD group across four consecutive 4-min sessions while controlling for multiple comparisons [53,54]. The session showing the strongest cluster-level effect was selected for subsequent analyses to capture the maximal tPBM-related effects. Source-level comparisons used the same nonparametric approach. EEG measures from the selected session were then analyzed using the same parametric statistical framework as the behavioral analyses

To investigate the relationship between tPBM-induced changes in EEG measures and working memory, cluster-based permutation Spearman correlations were performed between the differences (active minus sham) in EEG measures and in K scores. Values extracted from significant clusters were then entered into a commonality analysis to partition total explained variance (*R*^2^) into unique and common components [55]. All tests were two-tailed with an alpha level of 0.05.

## 3. Results

### 3.1. Safety and Behavioral Improvements of tPBM

To evaluate the tolerability of tPBM, self-reported experiences were collected after the intervention using questionnaires. No notable discomfort was reported, and the ratings did not differ between the active and sham conditions (*t*s_(27)_ ≤ 1.411, *p*s ≥ 0.174; Fig. 1D). Behaviorally, under the sham condition, participants with ADHD presented lower K scores than HCs did (independent-sample *t* test, *t*_(55)_ = –2.830, *p*_FDR_ = 0.020, Cohen’s *d* = –0.750; Fig. 1E). Following active tPBM, the K scores within the ADHD group significantly increased (*t*_(27)_ = 2.537, *p*_FDR_ = 0.026, Cohen’s *d* = 0.479; Fig. 1E), and the difference between the ADHD group and the HC group was no longer significant (*t*_(55)_ = –1.231, *p*_FDR_ = 0.224, Cohen’s *d* = –0.326; Fig. 1E).

### 3.2. Increased Alpha Power Following tPBM

To elucidate the neural dynamics of tPBM effects, we analyzed EEG data across four consecutive 4-min sessions spanning the stimulation period. To capture oscillatory signatures potentially related to E/I balance, we first examined changes in relative alpha power (Fig. 1C). At the sensor level, no clusters were found in the first and second sessions. In the third session, a significant increase in alpha power emerged under the active condition compared with the sham condition, with a frontal cluster forming ipsilateral to the stimulation site (cluster *p* = 0.045). In the fourth session, the effect became more robust and spatially extended, spreading posteriorly and additionally involving the contralateral frontal and posterior regions (cluster *p* = 0.002; Fig. 2A). Given that the strongest effects were observed in the fourth session, subsequent analyses focused on this time window to better capture peak tPBM-induced neural responses. Source-level analysis revealed increased alpha power in the active condition compared with the sham condition, primarily involving bilateral attention and frontoparietal systems, along with default mode and somatomotor networks, and extending to the left visual network (left: cluster *p* = 0.006; right: cluster *p* = 0.007; Fig. 2B).

**Fig. 2.**
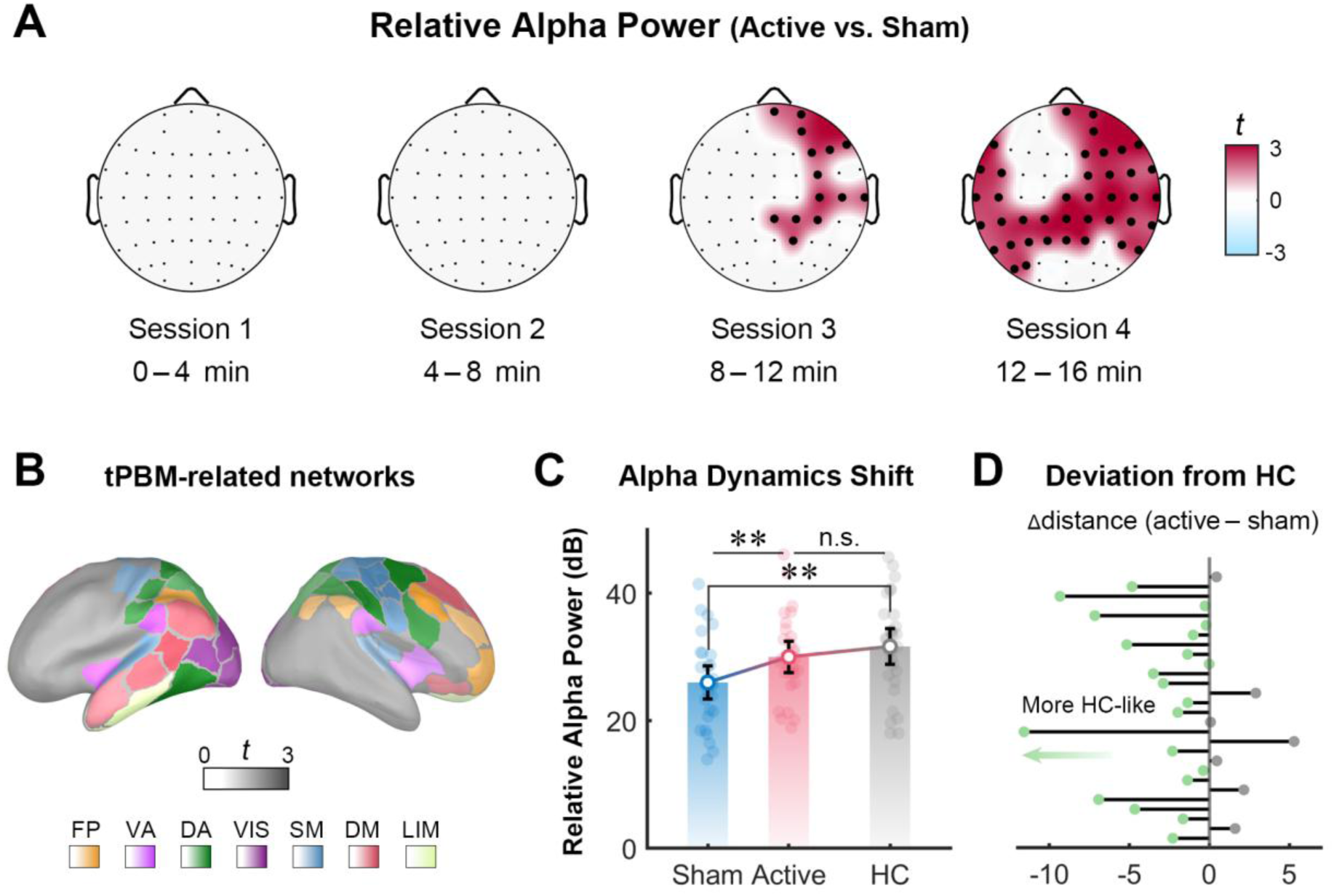
tPBM-related effects of relative alpha power. **(A)** Topographies showing tPBM effects (active vs. sham) on alpha power in the ADHD group across each 4-min session. Significant clusters are marked by bold dots (cluster-based permutation *t* test, *p* < 0.05, two-tailed). **(B)** Source-level tPBM-related networks for alpha power in the ADHD group, with color intensity reflecting the magnitude of change (cluster-based permutation *t* test, *p* < 0.05, two-tailed). FP, frontoparietal; VA, ventral attention; DA, dorsal attention; VIS, visual; SM, somatomotor; DM, default mode; LIM, limbic. **(C)** Cluster-derived alpha power in the ADHD group during session 4, comparing conditions (paired-sample *t* test), and between ADHD patients and HCs (independent-sample *t* test). The horizontal lines indicate the means ± standard errors of the means (SEMs), and the dots represent individual participants. n.s., no significant difference, \*\**p*_FDR_ < 0.01. **(D)** Deviation from HCs in alpha power. Subjectwise changes in deviation from HCs after active tPBM relative to sham in the ADHD group. The green dots denote responders, and the gray dots denote others.

A cluster-averaged analysis further revealed that, under the sham condition, ADHD participants exhibited lower alpha power than HCs did (*t*_(55)_ = –2.897, *p*_FDR_ = 0.005, Cohen’s *d* = –0.768). Compared with the sham condition, the tPBM condition significantly increased the alpha power in the active condition (*t*_(27)_ = 3.467, *p*_FDR_ = 0.002, Cohen’s *d* = 0.655). Importantly, the group difference between ADHD patients and HCs was no longer significant following active stimulation (*t*_(55)_ = –0.865, *p*_FDR_ = 0.391, Cohen’s *d* = –0.229; Fig. 2C). Moreover, at the individual level, approximately 75% of participants showed a smaller deviation from the HC-like distributed alpha activity pattern in the active condition than in the sham condition, suggesting a shift toward normalization of alpha activity in the ADHD group (Fig. 2D). Overall, these findings demonstrate a progressive spatial normalization of alpha activity following tPBM, reflecting potentially strengthened inhibitory regulation.

### 3.3. Steeper Aperiodic Exponent With tPBM

To probe broadband, nonoscillatory neural activity arising from E/I balance, the aperiodic exponent (1/f slope) was analyzed as a putative index of neural background activity (Fig. 1C). Similarly, no significant clusters were observed early in the stimulation period (cluster *p*s ≥ 0.148), with a significant effect emerging in the third session, where the exponent was greater in the active condition than in the sham condition, resulting in the formation of a central cluster (cluster *p* = 0.043). In the fourth session, the effect became stronger, encompassing the bilateral frontal and parieto-occipital regions (cluster *p* = 0.012; Fig. 3A). Source-level analysis further localized this effect, revealing a greater exponent in the active condition than in the sham condition, with effects on bilateral attention and frontoparietal networks, as well as the bilateral visual network and the left default mode network (left: cluster *p* = 0.036; right: cluster *p* = 0.043; Fig. 3B).

**Fig. 3.**
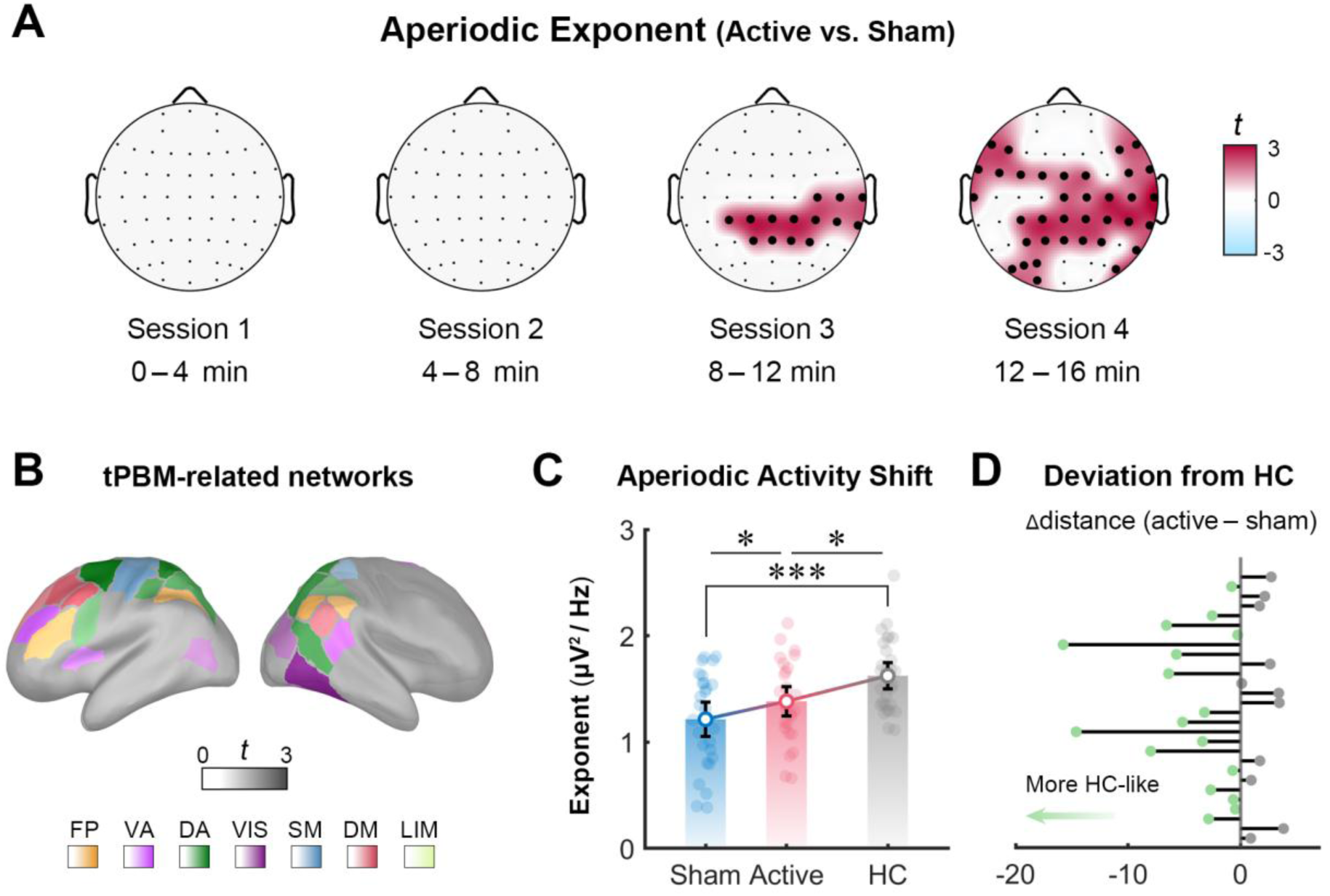
tPBM-induced modulation of the aperiodic exponent. **(A)** Topographical plots of tPBM-induced exponent changes (active vs. sham) in the ADHD group during the intervention. Significant clusters are indicated by bold dots (cluster-based permutation *t* test, *p* < 0.05, two-tailed). **(B)** Source-level tPBM-related networks for exponent in the ADHD group, with color intensity reflecting the magnitude of change (cluster-based permutation *t* test, *p* < 0.05, two-tailed). **(C)** Cluster-derived exponent in the ADHD group during session 4, comparing conditions (paired-sample *t* test), and between ADHD patients and HCs (independent-sample *t* test). The horizontal lines indicate the means ± SEMs, and the dots represent individual participants. \**p*_FDR_ < 0.05, \*\*\**p*_FDR_ < 0.001. **(D)** Deviation from HCs in the exponent. Subjectwise changes in deviation from HCs between active and sham tPBM conditions in the ADHD group. The green dots denote responders, and the gray dots denote others.

Extending this comparison to HCs, ADHD participants had significantly lower aperiodic exponent values than the HC pattern under the sham condition (*t*_(55)_ = –3.964, *p*_FDR_ = 0.001, Cohen’s *d* = –1.050). tPBM significantly increased the exponent in the active condition (*t*_(27)_ = 2.222, *p*_FDR_ = 0.035, Cohen’s *d* = 0.420), resulting in a reduced difference from the HC pattern, although a significant gap remained (*t*_(55)_ = –2.547, *p*_FDR_ = 0.021, Cohen’s *d* = –0.675; Fig. 3C). Furthermore, we examined whether tPBM induced convergence toward the HC-like distributed brain activity pattern at the individual level. A majority of participants (60.7%) exhibited this shift, indicating a movement toward a more typical neural organization and suggesting an amelioration of inhibitory dysfunction (Fig. 3D). Overall, these results indicate a temporally evolving steepening of the aperiodic spectral slope induced by tPBM, reflecting a progressive shift in E/I-related neural dynamics.

### 3.4. Higher Multiscale Entropy After tPBM

To further capture the complexity of neural dynamics shaped by E/I-related processes, MSE was analyzed across temporal scales (Fig. 1C). An increase in MSE, indicating greater neural complexity, was not observed in the first or second sessions (cluster *p*s ≥ 0.095). In the third session, however, a significant effect emerged, with higher MSE in the active condition than in the sham condition, resulting in the formation of a frontoparietal cluster at higher scales (cluster *p* = 0.031). This effect further extended to contralateral posterior regions, primarily across scales 11–20 (cluster *p* = 0.009; Fig. 4A). Source-level analysis further localized this effect, showing an increase in MSE within bilateral attention and frontoparietal networks, together with bilateral visual, default mode, and somatomotor networks (left: cluster *p* = 0.034; right: cluster *p* = 0.020; Fig. 4B), demonstrating more structured temporal organization of neural activity following tPBM, which may support more flexible neural processing.

**Fig. 4.**
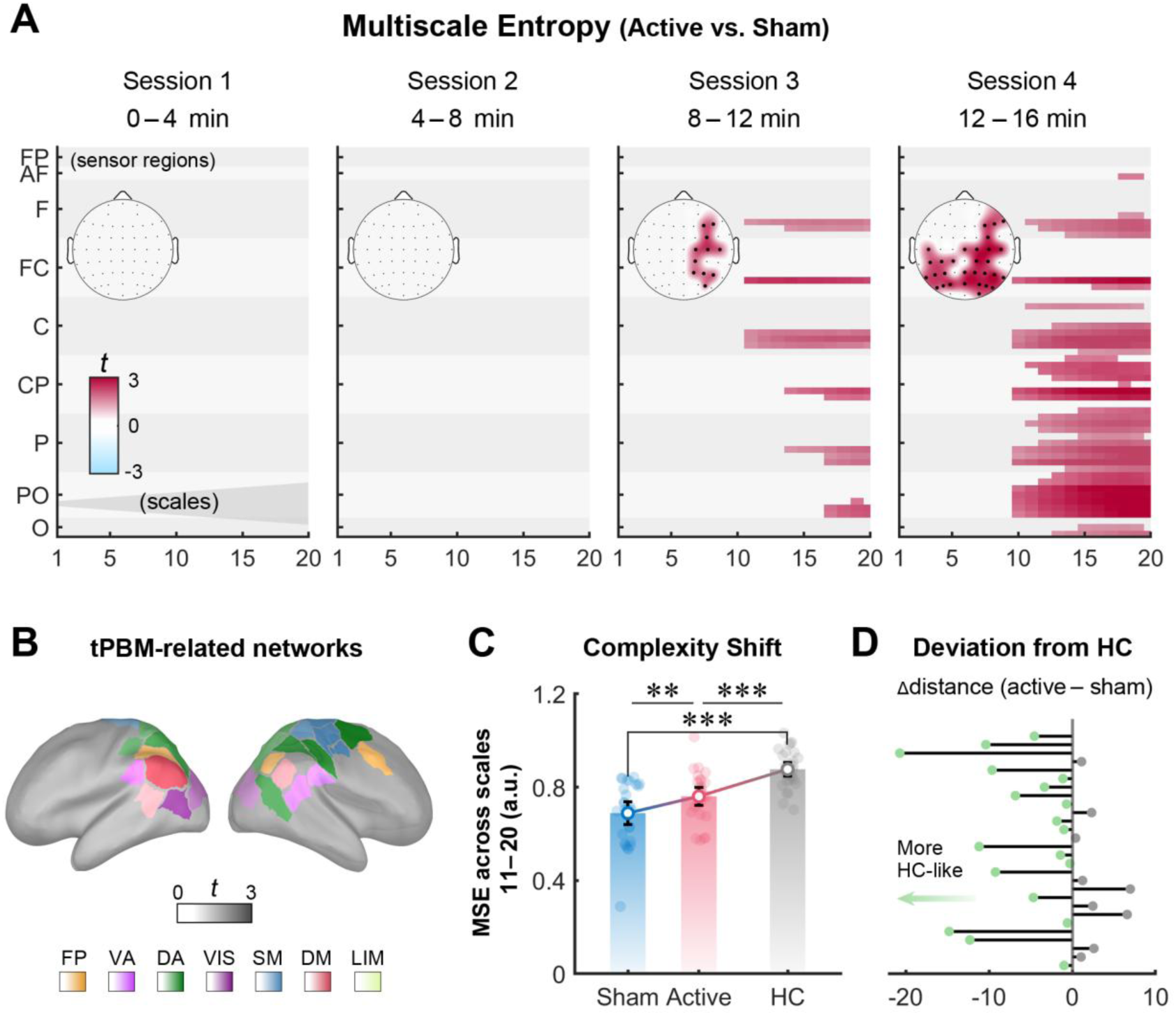
tPBM-associated changes in multiscale entropy. **(A)** tPBM-related changes in MSE (active vs. sham) across scales and sensor regions during intervention, with corresponding scalp distributions. Significant clusters in the scale–sensor space are outlined (cluster-based permutation *t* test, *p* < 0.05, two-tailed). **(B)** Source-level tPBM-related networks for MSE in the ADHD group, with color intensity reflecting the magnitude of change (cluster-based permutation *t* test, *p* < 0.05, two-tailed). **(C)** Cluster-derived MSE in the ADHD group during session 4, comparing conditions (paired-sample *t* test), and between ADHD patients and HCs (independent-sample *t* test). The horizontal lines indicate the means ± SEMs, and the dots represent individual participants. \*\**p*_FDR_ < 0.01, \*\*\**p*_FDR_ < 0.001. **(D)** Deviation from HCs in MSE. Subjectwise changes in deviation from HCs following active tPBM relative to sham in the ADHD group. The green dots indicate participants who improved, and gray dots represent others.

Cluster-averaged MSE values across sensor-level clusters and scales 11–20 revealed that compared with HCs under the sham condition, ADHD participants exhibited markedly reduced MSE (*t*_(55)_ = –6.556, *p*_FDR_ < 0.001, Cohen’s *d* = –1.737). tPBM significantly increased MSE in the active condition relative to that in the sham condition, indicating enhanced dynamic richness of neural activity following stimulation (*t*_(27)_ = 3.287, *p*_FDR_ = 0.003, Cohen’s *d* = 0.621). Although the MSE in the active condition still differed from that in the HC pattern (*t*_(55)_ = –4.755, *p*_FDR_ < 0.001, Cohen’s *d* = –1.260; Fig. 4C), there was a general trend toward a more typical neural state following tPBM. Furthermore, analyses at the individual level revealed that 67.9% of participants showed a shift toward a more typical distributed neural activity pattern after stimulation (Fig. 4D), indicating convergence toward HC-like neural organization.

### 3.5. Neural Predictors of Behavioral Improvement

To examine the association between tPBM-induced neural changes and working memory improvement, cluster-based permutation Spearman correlations were conducted during the fourth session between active–sham EEG changes and behavioral gains, identifying significant spatial clusters (Fig. 5A). Significant correlations at the sensor level were observed in posterior regions for alpha power (cluster *p* = 0.019), as well as in frontoparietal regions for the aperiodic exponent (cluster *p* = 0.029) and in frontal regions for MSE (cluster *p* = 0.029). Source-level analyses revealed a comparable spatial pattern, with alpha power primarily localized to bilateral dorsal attention and frontoparietal networks, along with bilateral visual and default mode networks (left: cluster *p* = 0.012; right: cluster *p* = 0.023). Exponent effects involved the left dorsal attention and default mode networks (left: cluster *p* = 0.032; right: cluster *p* = 0.153). MSE effects were associated with the right dorsal attention and frontoparietal networks (left: cluster *p* = 0.117; right: cluster *p* = 0.023). These behavior-related clusters partly overlapped with those showing overall EEG changes, suggesting dissociable spatial patterns of behaviorally relevant neural modulation. To further quantify these relationships, values extracted from significant sensor-level clusters were entered into Spearman correlation analyses. Greater increases in alpha power (ρ = 0.524, *p* = 0.005), a steeper 1/f slope (ρ = 0.479, *p* = 0.011), and a higher MSE (ρ = 0.385, *p* = 0.044) were significantly associated with greater improvements in working memory performance (Fig. 5A).

**Fig. 5.**
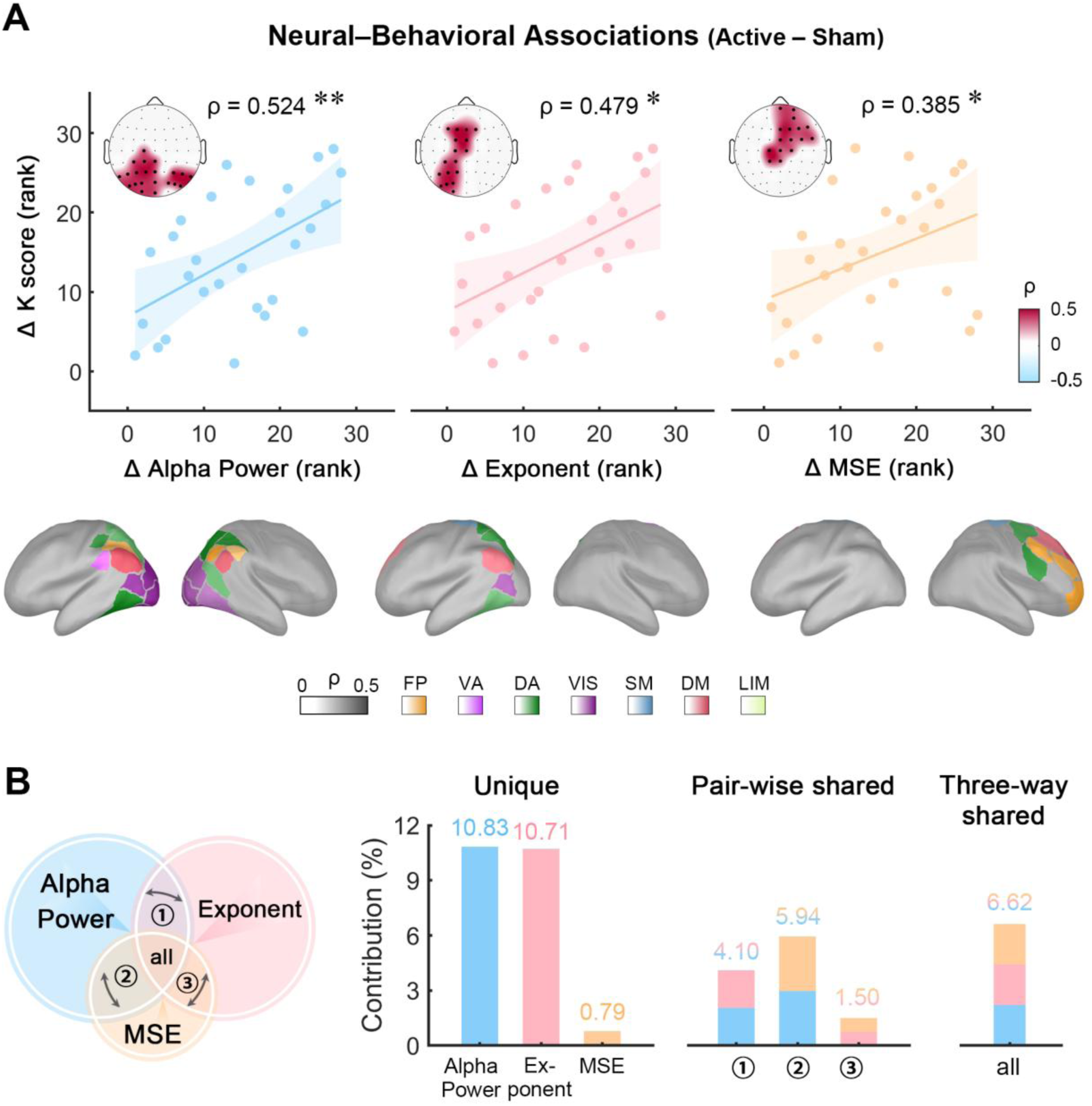
Associations between neural modulation and behavioral improvement. **(A)** Spearman correlations between tPBM-induced (active–sham) EEG changes during session 4 and working memory improvement, with cluster-extracted scatter plots and corresponding significant clusters marked by bold dots (*p* < 0.05, two-tailed). Higher ranks correspond to larger numerical values. Each dot represents an individual participant, and shaded areas denote 95% confidence intervals. The bottom panel further illustrates source-level networks associated with behavioral improvement, where darker colors indicate stronger correlations. \**p* < 0.05, \*\**p* < 0.01. **(B)** Commonality analysis quantifying the unique and shared contributions of the alpha power, exponent, and MSE to working memory improvement.

Commonality analysis was performed to determine the unique and shared contributions of each EEG measure to behavioral improvement (Fig. 5B). Together, the three EEG measures explained 40.49% of the variance in behavioral improvement. Unique contributions were observed for alpha power (10.83%), exponent (10.71%), and MSE (0.79%), alongside 6.62% of the shared variance among the neural metrics. In addition, alpha power and MSE contributed 5.94% of the explained variance. These results suggest that oscillatory and aperiodic dynamics provide relatively separable contributions, with their shared component across measures potentially reflecting a common E/I rebalancing process underlying tPBM-related working memory improvement.

## 4. Discussion

Accumulating evidence suggests that tPBM may enhance cognitive function, yet its neurophysiological mechanisms in ADHD remain poorly understood. Our findings show that tPBM enhances working memory in young adults with ADHD, increasing performance to a healthy level. This is accompanied by large-scale regulatory effects that stabilize cortical dynamics. Specifically, these effects include increased alpha power, a steeper aperiodic exponent, and improved neural complexity, indicating a coordinated reorganization of E/I balance. These effects extended beyond the stimulation site to the frontoparietal, attention, sensory, and default-mode networks, which is consistent with the partial normalization of atypical inhibitory dynamics. Critically, these neural changes tracked behavioral improvements in working memory, underscoring their functional relevance. Together, these results suggest that tPBM is a promising approach for restoring E/I balance and improving cognitive function in individuals with ADHD.

### 4.1. Working memory enhancement and neural reorganization following tPBM in ADHD

Working memory deficit is considered a central neurocognitive endophenotype of ADHD [2,3,56]. A growing body of evidence suggests that tPBM is effective at improving working memory. In healthy young adults, 12 minutes of 150 mW/cm^2^ stimulation over the rPFC is sufficient to enhance working memory [36], while similar benefits, including improved performance in the 3-back task, have also been observed in older adults following a single higher-irradiance (250 mW/cm^2^) intervention [57]. ADHD case reports also describe subjective improvements in memory and daily functioning after one-time stimulation, although evidence remains uncontrolled [35]. More recently, a behavioral study in adults with ADHD reported stronger effects in demanding tasks and individuals with poorer baseline performance, but comparisons among healthy individuals were lacking [37]. The present study used a randomized, double-blind, sham-controlled EEG design with HCs and demonstrated that tPBM (150 mW/cm², 16 min) enhances working memory in young adults with ADHD, resulting in performance close to that of healthy individuals.

With respect to neural activity, converging evidence suggests that ADHD is characterized by disrupted E/I balance and reduced inhibitory control within prefrontal and frontoparietal circuits [14,58,59]. Consistent with this framework, we observed altered oscillatory and aperiodic activity and reduced neural complexity, indicating inefficient neural regulation. Importantly, tPBM shifted these atypical neural signatures toward HC-like patterns, suggesting partial restoration of the E/I balance and improved neural efficiency [11]. Notably, all EEG indicators moved closer to the HC pattern, but only alpha power no longer showed a significant group difference, whereas aperiodic activity and MSE remained significantly different, indicating heterogeneous neural responses. Alpha oscillations may respond more rapidly to tPBM-induced metabolic effects, whereas aperiodic activity and neural complexity may reflect broader neurodevelopment-related alterations requiring longer or repeated intervention [60,61]. In the source space, ADHD-related abnormalities primarily involve attention and cognitive control networks [38,62]. Here, tPBM increased activity in frontoparietal executive, attention, and sensory regions, including visual and sensorimotor areas with reduced prefrontal coordination [63], as well as default-mode regions linked to internally oriented processes [64]. These results extend prior findings of widespread tPBM-related connectivity increases in healthy adults [65,66], demonstrating coordinated system-level modulation in ADHD.

### 4.2. The progressive effects of tPBM: temporal dynamics of rebalancing E/I in ADHD

Modulation emerged at intermediate stages and strengthened over time, suggesting a cumulative effect of tPBM rather than an immediate transient response. Previous work in healthy adults revealed that 11 min of rPFC 1064-nm tPBM (162 mW/cm²) progressively increased alpha oscillatory activity, with the strongest increase observed during 8–11 min of stimulation [67]. Another study using a higher irradiance (250 mW/cm²) similarly reported alpha-band modulation emerging during the second 4 min of stimulation and persisting into the post-stimulation period [68]. Consistent with these findings, alpha power, aperiodic activity, and MSE changes in the present ADHD study also became evident after approximately 8 min of stimulation (session 3), suggesting that the temporal dynamics of tPBM effects are broadly comparable between ADHD and healthy populations. Following near-infrared stimulation over the rPFC, neural changes were initially localized within right-lateralized frontoparietal networks and subsequently propagated to distributed cortical regions. This spatiotemporal pattern distinguishes tPBM from other neuromodulation techniques, such as transcranial direct current stimulation (tDCS) and repetitive transcranial magnetic stimulation (rTMS), which often induce more immediate but spatially constrained effects [69,70]. In contrast, tPBM may exert its influence through metabolic and mitochondrial pathways, enabling gradual, network-level reorganization and sustained rebalancing of E/I dynamics [71].

### 4.3. Shared and dissociable neural dynamics underlying tPBM-induced working memory enhancement

The present findings link tPBM-induced working memory improvement to specific neural dynamics in ADHD patients, providing a mechanistic account of its cognitive benefits. Our results indicate that associations with working memory improvement are partially aligned with the spatial distribution of overall tPBM-induced EEG changes, suggesting that widespread neural modulation does not map uniformly onto behavioral improvement but instead reflects a selective coupling between circuit-level changes and cognitive outcomes. Within these behaviorally relevant regions, increases in posterior alpha power, potentially reflecting enhanced inhibitory control and functional gating of task-irrelevant information [72,73], together with modulation of the aperiodic exponent, explained substantial variance in working memory improvement through both shared and independent contributions.

Specifically, these spectral features are not independent of broader signal properties. Low-frequency oscillatory power and the slope of the power spectral density are intrinsically linked to the temporal structure and irregularity of neural activity, which can be captured by measures of signal complexity such as MSE [55,74]. Consistent with this coupling, MSE contributed a smaller but still meaningful proportion of explained variance, largely shared with spectral changes. These findings align with those of prior works showing that higher neural entropy is associated with more efficient information processing and improved network efficiency [75,76]. Collectively, these results suggest that tPBM-related working memory enhancement primarily reflects increased neural flexibility, which may, at least in part, stem from coordinated shifts in oscillatory and aperiodic activity underlying the reorganization of E/I balance, with neural complexity indexing complementary integrative processes that support adaptive cognition.

### 4.4. The Therapeutic Potential of tPBM in ADHD

Current mainstream treatments for ADHD primarily include pharmacological approaches, such as methylphenidate and atomoxetine, as well as psychosocial interventions [77,78]. While effective, pharmacological treatments are often accompanied by side effects (e.g., sleep disturbances and appetite suppression) [79], and psychosocial interventions may face challenges in generalizability across contexts and individuals [80]. This highlights the need for interventions that are accessible and well tolerated, grounded in ADHD-related neural substrates, and compatible with existing treatment frameworks [81,82]. Noninvasive brain stimulation techniques, such as rTMS and tDCS, have shown some promise in modulating cognitive function in individuals with ADHD [83,84]. However, these approaches may induce auditory or somatosensory discomfort during stimulation, which could limit their acceptability, particularly in populations characterized by heightened sensory sensitivity [85,86]. Notably, no participants reported discomfort during tPBM in the present study, supporting its favorable safety and tolerability and potential as a more patient-friendly intervention [43].

The results of the current study provide evidence that tPBM can enhance core cognitive functions in ADHD patients via modulation of underlying neural balance, thus offering a promising avenue for functional recovery. Future studies should further examine task-related neural activity to provide more direct evidence of how tPBM facilitates adaptive behavior in ecologically valid contexts. In addition, longitudinal and dose‒response designs will be critical for characterizing the durability and optimization of tPBM effects, ultimately enabling more personalized intervention strategies. Finally, with appropriate ethical considerations, future work may extend the application of tPBM to broader developmental populations, facilitating wider clinical translation across age groups and related neurodevelopmental conditions.

## 5. Conclusion

This study provides converging evidence that tPBM ameliorates core cognitive deficits in young adults with ADHD. Behaviorally, 150 mW/cm^2^, 16 min tPBM enhanced working memory and resulted in performance closer to healthy levels. The effect of tPBM emerges progressively and extends from stimulation-targeted regions to large-scale brain networks, which is associated with increased alpha power, a steeper aperiodic exponent, and enhanced neural complexity, collectively shifting brain activity toward a more normative profile and ultimately contributing to behavioral gains in working memory. From the perspective of cortical homeostasis, these findings provide evidence that tPBM can rebalance atypical neural E/I patterns and alleviate core deficits in ADHD, offering a mechanistically supported basis for its application in neurotherapeutic interventions.

## CRediT authorship contribution statement

**Xuye Yuan:** Conceptualization, Methodology, Investigation, Data curation, Formal analysis, Visualization, Writing – original draft, Funding acquisition. **Yiyang Wang:** Conceptualization, Methodology, Investigation, Data curation, Formal analysis, Visualization, Writing – original draft. **Chen Dang:** Investigation, Data curation. **Hongyu Liu:** Investigation, Data curation. **Lili Yang:** Investigation, Data curation. **Dongwei Li:** Conceptualization, Methodology, Funding acquisition. **Li Sun:** Investigation, Writing – review & editing, Funding acquisition, Resources, Supervision. **Yan Song:** Conceptualization, Writing – review & editing, Funding acquisition, Resources, Supervision.

## Data Availability

The datasets and code generated during the current study are available in the OSF repository (https://osf.io/2wqf5/).

## Acknowledgments

The authors sincerely thank all the participants for their time and effort in this study.

