## Supplementary Fig. 1 for "Transcranial photobiomodulation rebalances cortical excitation/inhibition in young adults with attention deficit/hyperactivity disorder"

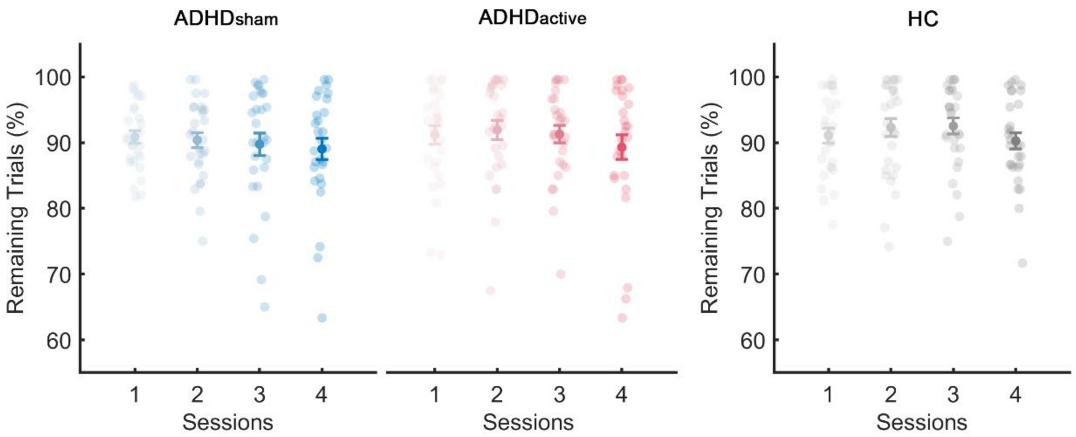


**Supplementary Fig. 1. Proportion of remaining trials for each condition and session.**

No significant differences were observed between the active and sham conditions in the ADHD group or across the four sessions. In addition, no significant differences were found between ADHD (active and sham conditions) patients and healthy controls (HCs) across sessions. A series of 2 × 4 repeated-measures ANOVAs was conducted to examine the effects of condition and session (ADHD within-group: condition effect $F_{(1, 27)} = 0.412$, $p = 0.526$, session effect $F_{(3, 81)} = 1.552$, $p = 0.208$, interaction $F_{(3, 81)} = 0.362$, $p = 0.780$; $\text{ADHD}_{\text{sham}}$ vs. HC: group effect $F_{(1, 55)} = 1.355$, $p = 0.249$, session effect $F_{(3, 165)} = 1.006$, $p = 0.392$, interaction $F_{(3, 165)} = 0.512$, $p = 0.675$; $\text{ADHD}_{\text{active}}$ vs. HC: group effect $F_{(1, 55)} = 0.171$, $p = 0.681$, session effect $F_{(1, 165)} = 1.996$, $p = 0.117$, interaction $F_{(1, 165)} = 0.163$, $p = 0.921$.
